# Estimation of the infection fatality rate and the total number of SARS-CoV-2 infections

**DOI:** 10.1101/2020.04.23.20077446

**Authors:** Carlos Hernandez-Suarez, Paolo Verme, Efren Murillo-Zamora

**Author notes:** Corresponding author: Email addresses (Carlos Hernandez-Suarez), (Paolo Verme), (Efren Murillo-Zamora).

## Abstract

We introduce a simple methodology to estimate the infection fatality rate (IFR) and from here the total number of infected with SARS-CoV-2. The virus has shown to be highly infectious and thus we based our method under the assumption that all members of a household with at least one confirmed case of COVID-19 should be infected, therefore we estimate the IFR using the number of secondary fatalities in households. The simplicity of the methodology allows for large sample sizes, since it requires minimal laboratory testing capabilities. We applied this methodology to a database of 3,232 confirmed cases in Mexico and arrived to an IFR estimate within the range reported in other studies.

## 1. Introduction

It is known that the immune response to SARS-CoV-2 may range from fully asymptomatic to exhibit mild or even severe responses that may cause death. Estimates of the probability of presenting a particular response are useful for prevention and attention purposes or even for building appropriate mathematical models that may provide some projections at the population level, specially to analyze the evolution of the immune population with the purpose of economic recovery. These estimates are particularly important to estimate the total number of infections by expanding the fraction of observed in some category, for the instance the number of hospitalized persons or the number of deaths.

Let *p* be the probability that an individual will die given that it is infected with SARS-CoV-2, that is, *p* is the infection fatality rate (IFR). If 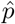 is and estimate of *p* then we can build an estimate of the number of infections per every death as 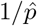. If the total number of deaths M is known, one can estimate the total number of infections with 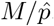.

There are current estimates of the probability of showing a specific reaction to infection, for instance, being asymptomatic, presenting mild or severe symptoms [1, 2, 3, 4], but their statistical properties are unknown. A possible design that would allow to estimate *p* is random screening for infection or antibodies, and categorizing the response of infected or already immune individuals. Some of these studies have been recently released for Iceland [5] and there are ongoing studies in other countries.

Here we suggest a simple study design based on the number of deaths observed in households with at least one confirmed case of COVID-19.

## Methodology

Let’s define an *effective contact* or *contact* for short as any act between an infectious and a susceptible individual that would result in the infection of the susceptible [6]. Let’s suppose that have *n* individuals that we know had a *contact*. Then and estimate of the IFR is 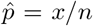 where *x* is then number of observed deaths among the *n* individuals.

From here, the importance of finding individuals that we know had a *contact*. But these individuals may easy to find: several studies have suggested that household transmission as well as familial transmission is very high [7, 8, 9, 10, 11, 12, 13] or even in offices for relative short interactions [14]. Therefore, if we are willing to concede that all the members of a household with a diagnosed individual had a *contact* with the initial infected in the household, the fraction of deaths among the remaining members of the household is an estimate of *p*. It is possible to pool data from several households to obtain a better estimate. In what follows, we formalize this estimate.

Suppose that we have a confirmed case of COVID-19. This confirmed case can lead us to household *j* with *n_j_* members in total. Define the individual that led us to a household as the *index case* (not necessarily the first case in a household). Assume that:

i. The remaining *n_j_* − 1 members of household are infected with probability 1.
ii. Once infected, the response to infection of each of the *n_j_* − 1 individuals are independent, that is, the number of deaths among the remaining susceptible members in a household follows a binomial distribution with parameters *n_j_* − 1 and *p*.

Observe that (i) implies that when two or more individuals are infected in the household, the probability that any one of the remaining susceptible will be infected is not increased. Also, it implies that all infected individuals are equally infectious, regardless of their symptomatic response to infection. Observe also that by excluding the *index case* of each household we avoid any bias.

### Estimation of IFR and the total number of infections

Suppose a sample of m confirmed individuals led us to *m* households of size *n_j_*, *j* = 1, 2, 3,…, *m*. Let 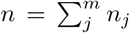 be the sum of all members in all households in the sample. Let *x_j_* be the number of deaths in household *j* (excluding all possible deaths of *index cases*) and let 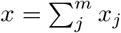. The estimate of *p*, the IFR measured at the household level is *x_j_*/(*n_j_* − 1). Using all households data in the sample, the estimate of *p* is:

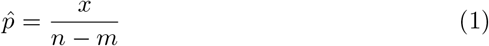

with variance 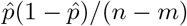.

With one further assumption, one can estimate the number of infections for the total population from these same data. If we assume that the number of COVID-19 deaths recorded includes all deaths from COVID-19, we can simply estimate the number of infected people in the population by expanding the fraction of infected people estimated from the sample of observed households. This should provide a simple but statistically sound estimate of the total number of infected people in the population.

The estimate of the total number of infections per death is about 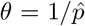. The approximate variance of 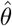 is:

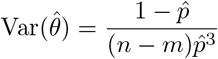

Let *M* be the total number of deaths from COVID-19 in the population, the estimate of the total number of infected individuals in the population, *N* is:

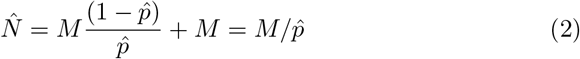

with approximate variance:

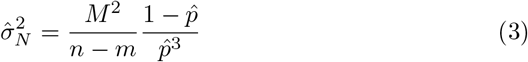

It is important to stress that our model does not assume that all infections among the remaining *n_j_* − 1 members of the household were caused by the same individual. In fact, our approach only requires that the remaining *n_j_* − 1 individuals in the household have had enough infectious pressure to guarantee they are infected. Thus, it works even if one or more infections among the members in a household were caused outside the household.

## Example

In this example we build an approximation to (1) using a database from Mexico’s IMSS (Instituto Mexicano de Seguro Social), the Mexican Institute for Social Insurance.

The database has 9939 confirmed SARS-CoV-2 cases from March 2 to May 4, 2020. In an attempt to consider only households with final outcomes we excluded cases with symptoms onset in the last 21 days, that is, we considered only cases from March 2 to April 19, 2020. The final dataset has 3232 cases.

We grouped the cases in households. If there were more than one case in a household, we only considered the household if all cases were already solved as deaths or recoveries. In every household with more than one case we consider the index case as the individual with the earliest symptom onset and counted the number of deaths among the remaining members of the house. From the final set of 3193 households, there were 3185 with no deaths among the remaining members of the household and 8 houses with one additional death. The mean age of this final set was 46.0 years with a standard deviation of 14.79 years with median 45 years. From these, there were 57.4 % males and 42.6 % females. In this set, 37 % were at least 50 years old.

The total number of households was *m* = 3193 and there were a total of *x* = 8 deaths. Since the total number of individuals in all households in the sample (*n*) is not known, we vary the average household size in the sample (*μ*) to calculate *n* = *mμ* and estimate 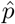 using (1). The results are summarized in Figure 1.

**Figure 1:**
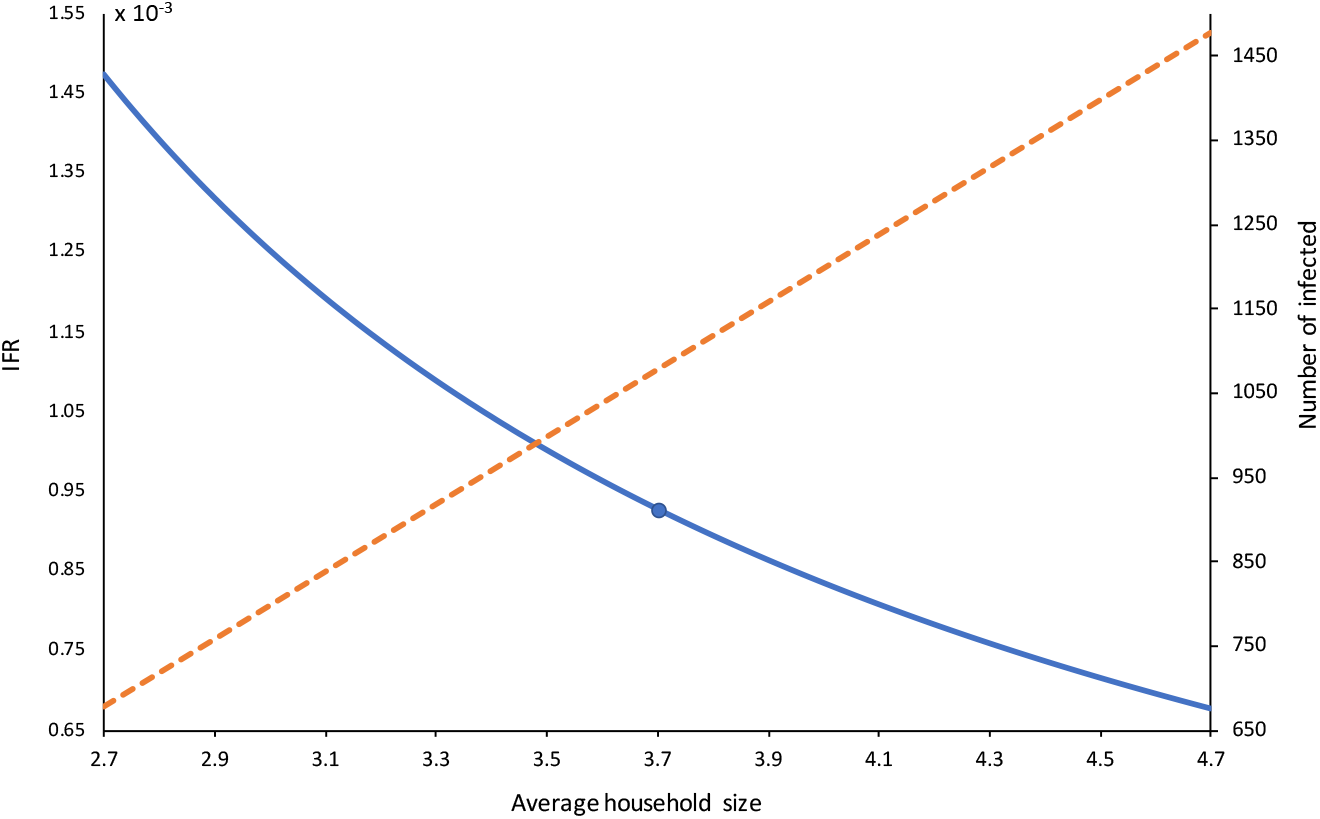
Plot of the average household size vs estimated IFR and K=1/IFR^−1^, the ratio of the total infected to deaths. The point marks the IFR at the average family size for Mexico, 3.7 At this average, IFR= 0.00092 and *K* = 1077.

## Discussion

First we must mention that our goal here is not to provide precise estimates of *p* for Mexico since the total number of individuals in all households is not known and we used an approximation according the average household size. Our goal is to illustrate a simple methodology to estimate the true number of infections in a population using available information on confirmed individuals.

Our estimate from the IMSS data at the average household size *μ* = 3.7 is *p =* 0.00092, which is in the range 0.001 – 0.003 reported in [15] and [16].

Our method is simple enough to be applied in countries with relatively few tracking capabilities. All it is needed is a list of households with at least one confirmed case of COVID-19 (a sample may suffice) with the total number of members in the household and the number of deaths for COVID-19 in each household. The precision of estimate (1) depends on the sample size *m*, and the precision of estimate (2) depends in addition on how good is our estimate of the actual number of deaths from COVID-19 to date. Overall, the precision will depend on our ability to diagnose COVID-19 related deaths.

Assumption (i) is central for this proposal, but there is a way to avoid it although clearly at a larger economic cost: this consists in testing all the members of the household of a confirmed case. The estimate (1) can still be applied using only data of confirmed cases, but now *x* is the number of deaths among all confirmed cases in all households (excluding the *infected zero*) and *n* the total number of confirmed cases in all households (including the *infected zero*).

In a following step, we can obtain the same probabilities for the whole population of positive cases by matching the household sample of tested households with households in the census. In other words, we only need to make sure that the sample of households retained from the interviews is representative of the national sample of households. This can be done, *ex-ante* with a sample of available infected households or, if this information is not available, *ex-post* by matching the interviewed sample of households with the national census of households. Something that can be done with matching or machine learning methods. This provides the distribution of cases between any categorization of symptoms for the population of infected people in a population. A direct approach from stratified sampling may use some demographic knowledge of the population which would allow us to weight for differential response to the infection. Suppose that we classify a population in K categories (e.g., age) at relative frequencies *f_i_*. Let *x*^(^*^i^*^)^ and *n*^(^*^i^*^)^ be respectively the total number of deaths and total number of individuals in category *i* in all households in the sample of size *m*, then a better estimate of *p* would be:

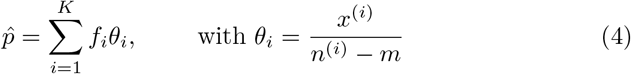

with variance

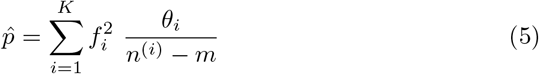

This 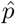 must be plugged in (2), with variance (3). We can divide then population in Mexico in two categories: age ≤ 50 years and age > 50 years, at respective proportions *f*_1_ = 0.9 and *f*_2_ =0.1 [17]. The IFR in the first category was 0.002 and in the second 0.0052. From (4) we have 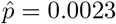 for the whole population, the weighted estimate suggests the number of total infected is about 400 times larger than the number of deaths.

One of the most important sources of bias in this method, is that some observations may be censored. Perhaps death has not occurred yet in a given household and thus the probability of death is underestimated. In our analysis of IMSS data, we tried to control this by using only data where the onset of symptoms was at least 21 days old so that the outcome is very likely observed, but in principle, we should use households were there is enough evidence to believe that we can observe final outcomes.

## Data Availability

No data made available

## Conflict of interest

Authors declare no conflict of interest.

## Funding

This work is part of the program “Building the Evidence on Protracted Forced Displacement: A Multi-Stakeholder Partnership”. The program is funded by UK aid from the United Kingdom’s Department for International Development (DFID), it is managed by the World Bank Group (WBG) and was established in partnership with the United Nations High Commissioner for Refugees (UNHCR). The scope of the program is to expand the global knowledge on forced displacement by funding quality research and disseminating results for the use of practitioners and policy makers. This work does not necessarily reflect the views of DFID, the WBG or UNHCR. This study had approval R-2020-601-07 by the Health Research Ethics Committee (601) of the IMSS.

## Notes

### Competing Interest Statement

The authors have declared no competing interest.

